# Noncoding de novo mutations in *SCN2A* are associated with autism spectrum disorders

**DOI:** 10.1101/2024.05.05.24306908

**Authors:** Yuan Zhang, Mian Umair Ahsan, Kai Wang

## Abstract

Previous genetic studies in ASD identified hundreds of high-confidence ASD genes enriched with likely deleterious protein-coding de novo mutations (DNMs). Multiple studies also demonstrated that DNMs in the non-coding genome can contribute to ASD risk. However, identification of individual risk genes enriched with noncoding DNMs has remained largely unexplored. We analyzed two datasets with over 5000 ASD families to assess the contribution of noncoding DNMs. We used two methods to assess statistical significance for noncoding DNMs: a point-based test that analyzes sites that are likely functional, and a segment-based test that analyzes 1kb genomic segments with segment-specific background mutation rates. We found that coding and noncoding DNMs in *SCA2A* are associated with ASD risk. Further application of these approaches on large-scale whole genome sequencing data will aid in identifying additional candidates ASD risk genes.

## Introduction

Autism spectrum disorder (ASD) is a neurodevelopmental disorder characterized by impaired social interaction and communication, and restrictive interests or repetitive behaviors^1–4^. ASD begins before the age of 3 years and can last throughout a person’s life, though symptoms may improve over time^5^. A recently review conducted by Tomoya et al. reported that ASD affects approximately 2.3% of children aged 8 years and approximated 2.2% of adults in the US^6^. Additionally, it has a strong male bias, with boys being affected nearly 4 times more common than girls among children aged 8 years^7^. ASD is often accompanied by other psychiatric disorders, such as intellectual disability, attention-deficit hyperactivity disorder, as well as anxiety, depression, schizophrenia, irritability, and aggression^1,8^. The neural mechanisms underlying impairment observed in ASD remain unknown, though recent work from genetic, imaging, molecular biology, and gross anatomy investigations has provided important insights.

ASD exhibits extensive clinical and genetic heterogeneity with high heritability^9–11^, and previous studies estimated that common variants explain ∼50% of the heritability^9^, with a few genome-wide significant loci identified^12^. Despite the evidence of a significant role for common variants in ASD risk, rare genetic variation (MAF<1%) confers higher individual risk^13,14^. The rare inherited variants account for a portion of the heritability^2,9^, while *de novo* mutations (DNMs) identified from parent–offspring trios are the underlying cause for many cases of ASD, explain additional proportions of the overall liability^9^. Many previous genetic studies in ASD focused on DNMs have identified hundreds of high-confidence ASD genes enriched with likely deleterious protein-coding DNMs^15–22^, and most of them are also implicated in other neurodevelopmental disorders (NDDs)^21,23,24^. A large-scale exome sequencing study, using an enhanced analytical framework to integrate *de novo* and inherited rare coding variants, identified 102 putative ASD-associated genes (e.g., *CHD8*, *SCN2A*, *ADNP*)^21^. Moreover, Zhou *et al*. performed a two-stage analysis of rare *de novo* and inherited coding variants and identified 60 genes with exome-wide significance (p < 2.5×10^−6^), including five new risk genes (*NAV3*, *ITSN1*, *MARK2*, *SCAF1*, and *HNRNPUL2*)^22^. Nevertheless, statistical modeling suggests that there are ∼1,000 genes with DNMs associated with ASD^14,25^. Additionally, previous genetic studies also demonstrated that DNMs in the non-coding genome can contribute to ASD risk^26–30^. Yuen et al. performed whole-genome sequencing of 200 ASD parent–child trios to characterize DNMs and found a significant enrichment of predicted damaging DNMs in ASD cases, of which 15.6% were non-coding^27^.

Meanwhile, the authors revealed that non-coding elements most enriched for DNM were untranslated regions of genes, regulatory sequences involved in exon-skipping and DNase I hypersensitive regions. Kim *et al*. generated 813 whole-genome sequences from 242 Korean simplex families and found that target genes, including *ARHGEF2*, *BACE1*, *CDK5RAP2*, *CTNNA2*, *GRB10*, *IKZF1*, and *PDE3B*, affected by the non-coding DNMs in chromatin interactions led to early neurodevelopmental disruption implicated in ASD risk^30^. However, identification of the individual risk genes enriched by noncoding DNMs has remained elusive, and the pathogenic effects of noncoding DNMs related to ASD are still poorly understood. Therefore, identifying noncoding DNMs that regulate gene function could provide important insights into ASD pathophysiology, which may have implications for targeted therapeutics.

Simons Foundation Powering Autism Research for Knowledge (SPARK)^31^ is an autism research initiative that aims to recruit and retain a community of 50,000 autistic individuals and their family members to advance understanding of the genetic basis of ASD. As of March 2023, SPARK generated whole-genome sequencing data for 12,519 individuals from over 3000 families, offering the opportunity to assay the contribution of noncoding DNMs for ASD risk. Here, we analyzed whole-genome sequencing data to identify both coding and noncoding DNMs in 3509 ASD trios comprised of affected probands and unaffected parents and in 2218 unaffected sibling-parent trios from the SPARK cohort. To evaluate genes significantly enriched for noncoding DNMs, we used two methods to assess statistical significance: a point-based test that analyzes sites with a Combined Annotation Dependent Depletion (CADD)^32^ score ≥15, and a segment-based test that analyzes 1kb genomic segments with segment-specific background mutation rates (inferred from expected rare mutations in Gnocchi genome constraint scores)^33^. Additionally, we replicated our findings in the Simons Simplex Collection (SSC)^34^ cohort with 6383 individuals from 2274 families.

## Results

### Cohort characteristics and study workflow

Our custom DNMs analytic pipeline was shown in **Fig 1**. A total of 3508 affected probands and 2218 unaffected sibling control from 3357 family trios were included in SPARK cohort (March 2023 release). Over 65% of the families are quartets. The mean (standard deviation) age of the SPARK cohort in this analysis was 9.0 years (5.9 years) for affected probands, 7.9 (4.5) for unaffected siblings, and 40.1 (8.4) for parents. The breakdown of sex in the full SPARK cohort was 58.2% male and 41.8% female, while among ASD cases, the breakdown was 79.7% male and 20.3% female. The characteristics of the individuals included in our analysis was shown in **Table 1**. Additionally, the SSC cohort, comprising of 6383 individuals (including 2274 affected probands and 1835 unaffected siblings) from 2274 families (over 80% were quartets), were applied to replicate the *de novo* variants analysis.

**Fig 1.**
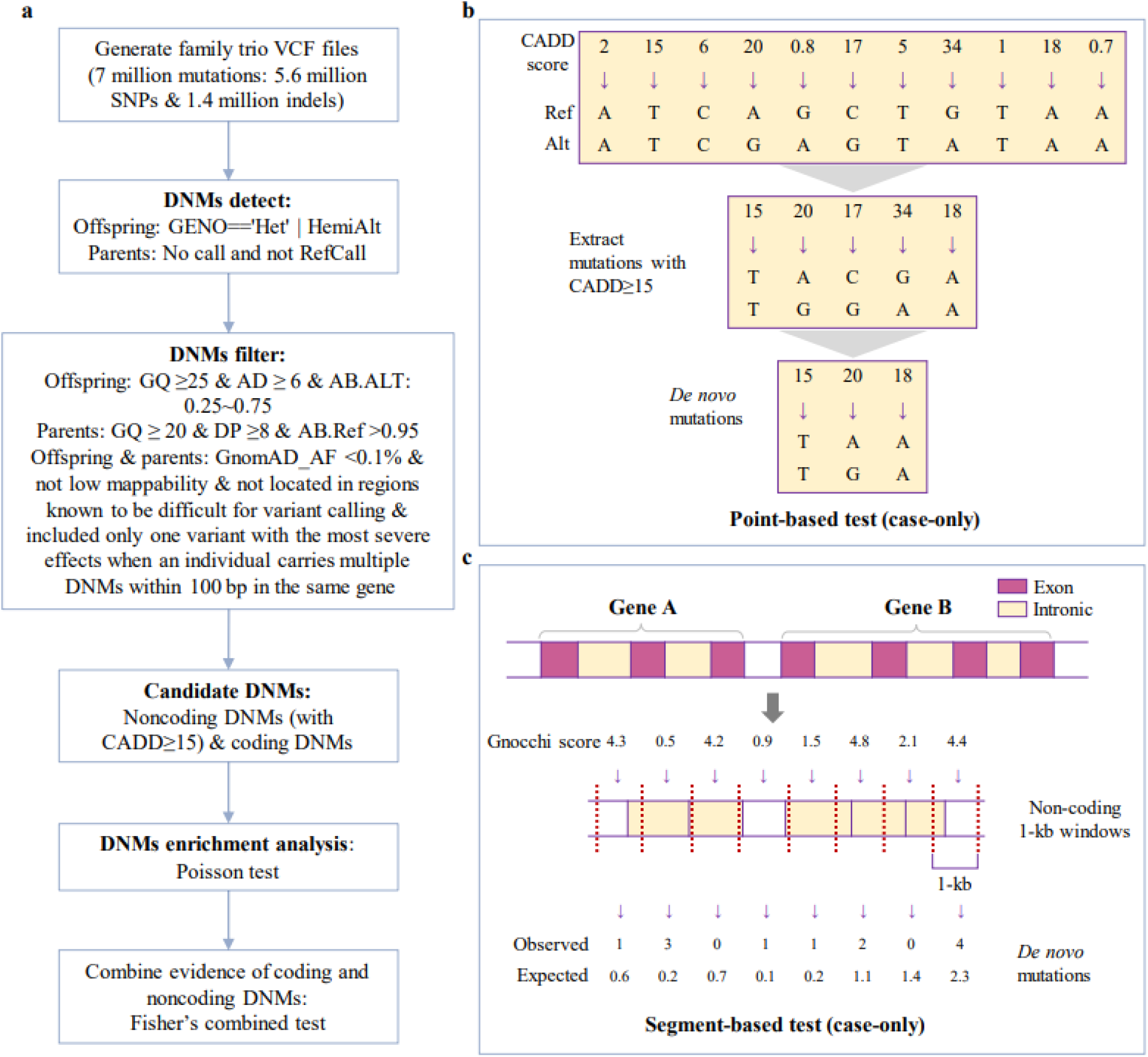
Analysis workflow. **(a)** DNMs analytic pipeline. For noncoding DNMs, this study used two methods to assess statistical significance **(b)** point-based test that analyzes sites with a Combined Annotation Dependent Depletion (CADD) score ≥15, and **(c)** segment-based test that uses Gnocchi genome constraint scores in 1kb genomic segments to infer background mutation rates.

**Table 1.** Characteristics of individuals included in the SPARK cohort.

| Characteristic | Autism case<br>(n=3508) | Sibling control<br>(n=2188) | Parents<br>(n=6714) |
| --- | --- | --- | --- |
| Age at enrollment in years, mean (SD) | 9.0 (5.9) | 7.9 (4.5) | 40.1 (8.4) |
| Sex at birth, n (%) |  |  |  |
| Male | 2796 (79.7) | 1069 (48.9) | 3358 (50.0) |
| Female | 712 (20.3) | 1119 (51.1) | 3356 (50.0) |
| Genetically inferred race/ethnicity, n (%) |  |  |  |
| White | 2648 (75.48) | 1656 (75.69) | 5121 (76.28) |
| African American | 168 (4.79) | 96 (4.39) | 325 (4.84) |
| Asian | 195 (5.56) | 123 (5.62) | 384 (5.72) |
| Hispanic | 382 (10.89) | 240 (10.97) | 740 (11.02) |
| Other | 115 (3.28) | 73 (3.34) | 143 (2.13) |

### Identification of coding DNMs for ASD

Through our custom pipeline designed for *de novo* analysis, we identified an average of 1.27 coding DNMs per affected offspring and 1.21 per unaffected siblings in the final call set (**Table 2**). In the ASD case group, 9.5% (424/4,446) of the coding DNMs were classified as likely gene disrupting (LGD), and 61.4% (2,732/4,446) were characterized as missense. While in the unaffected sibling control group, 7.4% (196/2,650) of the DNMs were classified as LGD, and 62.6% (1,660/2,650) were missense mutations. The number of coding DNMs observed per trio approximately follows Poisson distribution (**Fig 2**). SSC was used to replicated DNM analysis. The average number of coding DNMs is 1.38 for probands and 1.30 for unaffected siblings. Additionally, 9.3% of the coding DNMs were classified as LGD in the ASD group, while 6.1% of the coding DNMs were classified as such in the unaffected sibling control group. The distribution of coding DNMs from SSC was shown in **Fig S2**.

**Fig 2.**
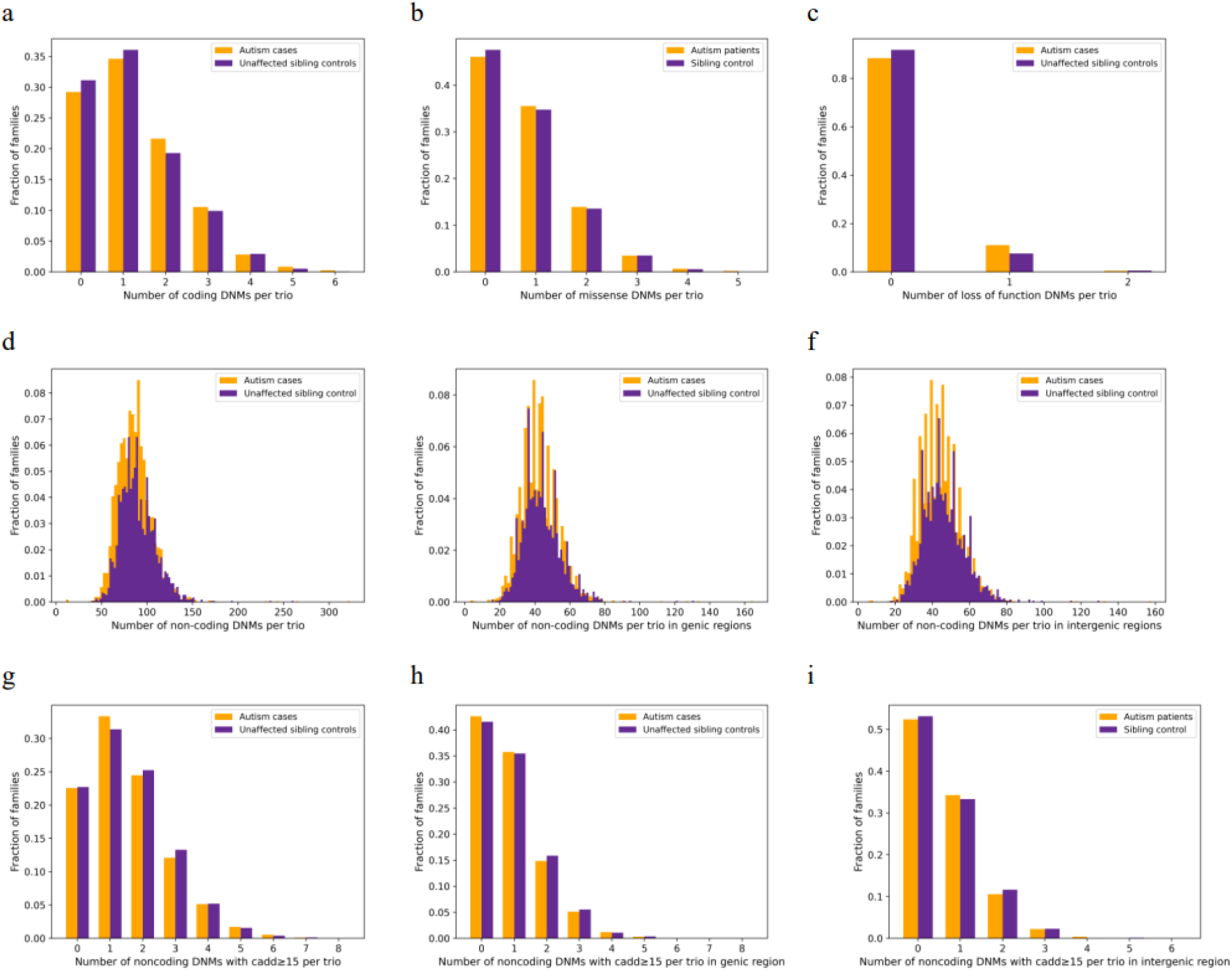
Distribution of DNMs in SPARK. **(a)** distribution of coding DNMs. **(b)** distribution of missense DNMs. **(c)** distribution of loss of function DNMs. **(d)** distribution of noncoding DNMs. **(e)** distribution of generic noncoding DNMs. **(f)** distribution of intergenic noncoding DNMs. **(g)** distribution of noncoding DNMs have a CADD score ≥15. **(h)** distribution of generic noncoding DNMs have a CADD score ≥15. **(i)** distribution of intergenic noncoding DNMs have a CADD score ≥15.

**Table 2.** Summary of *de novo* mutations (DNMs) identified from whole genome sequencing.

| Categories | SPARK cohort |  | SSC cohort |  |
| --- | --- | --- | --- | --- |
|  | ASD cases<br>(n=3508) | Unaffected<br>sibling controls<br>(n=2188) | ASD cases<br>(n=2274) | Unaffected<br>Sibling controls<br>(n=1835) |
| **Coding DNMs** | 4446 | 2650 | 3143 | 2390 |
| Likely gene disrupting <sup>a</sup> | 424 | 196 | 292 | 146 |
| Missense | 2732 | 1660 | 1925 | 1501 |
| Coding DNMs per trio | 1.27 | 1.21 | 1.38 | 1.30 |
| **Non-coding DNMs** | 302,603 | 196,898 | 209,396 | 171,258 |
| With CADD $\geq 15$ | 5343 | 3424 | 3645 | 2920 |
| Genic | 148,716 | 95,875 | 99,960 | 80,981 |
| With CADD $\geq 15$ | 3090 | 2010 | 2119 | 1703 |
| Intergenic | 153,887 | 101,023 | 109,436 | 90,277 |
| With CADD $\geq 15$ | 2253 | 1414 | 1526 | 1217 |
| Noncoding DNMs per trio | 86.3 | 90.0 | 92.1 | 93.2 |

<sup>a</sup> Likely gene disrupting mutations including frameshift deletion, frameshift insertion, startloss, stopgain, stoploss and splicing.

We applied case-control comparison to evaluate the burden of coding DNMs between ASD cases and unaffected sibling controls. Fold change was calculated as the ratio between affected offspring and unaffected sibling controls. We identified nine genes with p <0.1 (DNM count ≥ 3, fold change>1), including *SCN2A*, *BRD4*, *GRIN2B*, *WDFY3*, *CHD2*, *SPEN*, *CACNA1I*, *ADNP*, and *KDM6B*. Notably, *SCN2A* had a p=4.81×10^−3^. When replicating these gene in SSC, only *SCN2A* reached p<0.1 (p=0.052). Additionally, *CHD8* achieved the lowest p-value (p=0.0158) in SSC (**Table S3**).

We identified genes significantly enriched for coding DNMs by comparing the observed gene-wise DNM count to that of expected based on a model of gene-specific mutation rates ^35^ (case-only test). P-values for case-only tests are calculated from Poisson distribution using counts of observed/expected DNMs in ASD cases. The Quantile-Quantile plot (Q-Q plot) of p-values for point-based test was shown in **Fig 3**. We identified 120 genes that harbor an excess of coding DNMs (*p* <0.05 by case-only test, DNM count ≥ 3; **Table S3**). Among them, *SCN2A* and *CCDC168* with the excess of coding DNMs met Bonferroni corrected significance (*p* < 2.5×10^−6^ by case-only test). In parallel, we identified 76 genes with an excess of coding DNMs (*P* <0.05 by case-only test, DNM count ≥ 3) from the SSC cohort, three of which (*CHD8, AHNAK2*, and *FLG2)* reached the Bonferroni corrected significance (*p* < 2.5×10^−6^ by case-only test). The overlapping genes that are enriched for coding DNMs between SPARK and SSC cohorts includes *SCN2A*, *ANKRD11*, *GRIN2B*, *KDM6B*, *ARID1B*, *SPEN*, *PTPRF*, and *DNMT3A*, with *PTPRF* being a previously unreported candidate gene (*p* <0.05 in both cohorts, DNM count ≥ 3).

**Fig 3.**
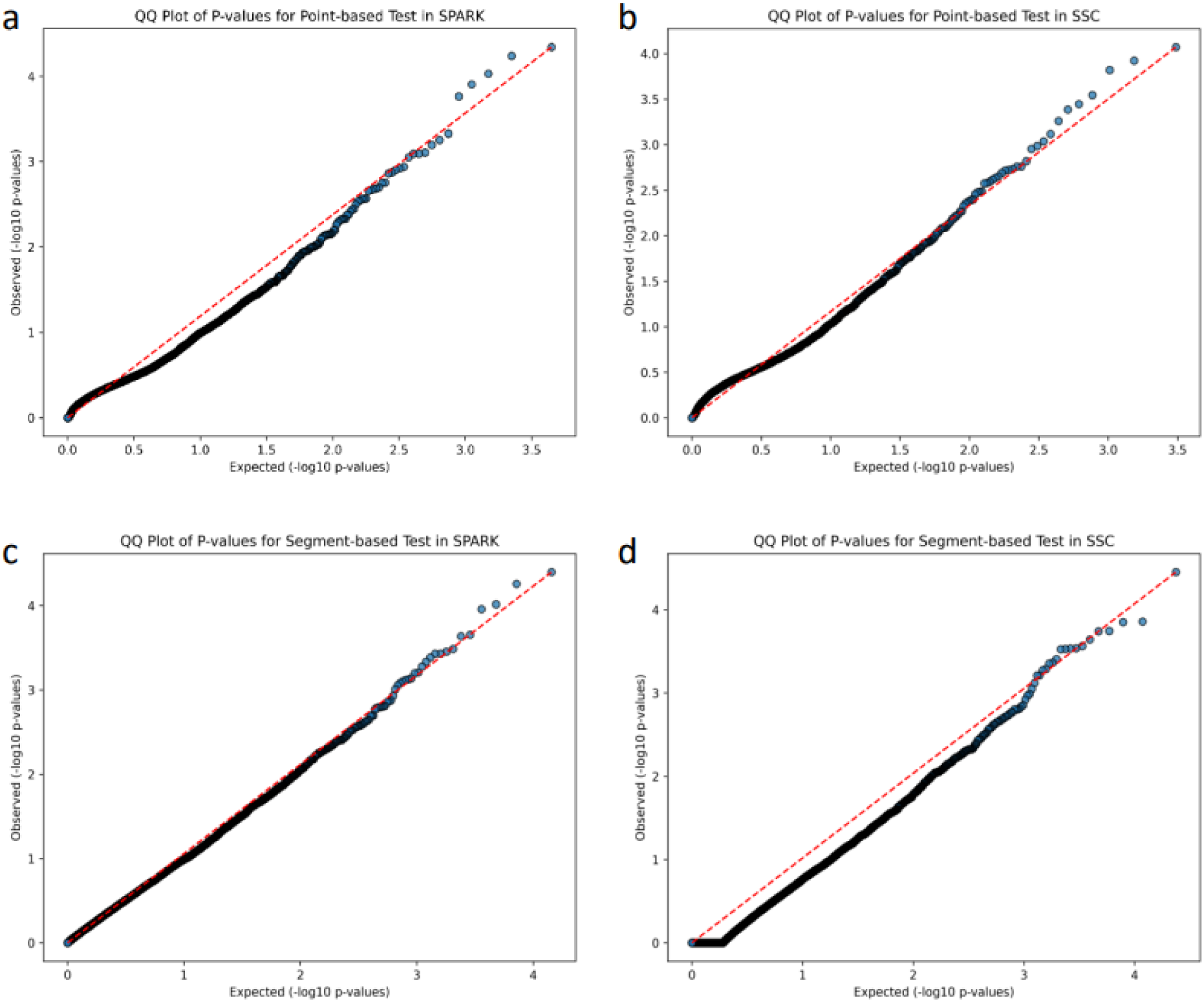
Q-Q Plot of P-values from point-based and segment-based tests. (**a**) Q-Q Plot of P-values for point-based test in SPARK. (**b**) Q-Q Plot of P-values for point-based test in SSC. (**c**) Q-Q Plot of P-values for segment-based test in SPARK. (**d**) Q-Q Plot of P-values for segment-based test in SSC.

We further classified coding DNMs as missense and LGD DNMs. We compared the observed missense/LGD DNM count to that of expected and identified 34 genes that were enriched for missense DNMs (*p* <0.05 by case-only test, DNM count ≥ 3) and 11 genes that were enriched for LGD DNMs (*p* <0.05 by case-only test, DNM count ≥ 3). Notably, four genes (*SCN2A*, *BRSK2*, *CCDC168*, *ADNP*, and *PRICKLE2*) with LGD DNMs met Bonferroni corrected significance (*p* < 2.5×10^−6^ by case-only test). For SSC, 23 genes were enriched for missense DNMs (*p* <0.05 by case-only test, DNM count ≥ 3) and 3 genes were enriched for LGD DNMs (*p* <0.05 by case-only test, DNM count ≥ 3). *CHD8* with 6 LGD DNMs met Bonferroni corrected significance (*p* < 2.5×10^−6^ by case-only test).

### Analysis of genes harboring noncoding DNMs associated with ASD

Besides coding DNMs, we also utilized our custom pipeline to identify noncoding DNMs. By comparing each affected and unaffected offspring to their parents, 302,603 noncoding DNMs (148,716 in genic regions and 153,887 in intergenic regions) were identified from 3508 affected offspring trios, while 196,898 DNMs (95,875 in genic regions and 101,023 in intergenic regions) were identified from 2218 unaffected sibling trios in the SPARK cohort. The average of noncoding DNMs is 86.3 per proband and 90.0 per unaffected sibling.

We initially used a point-based test to assess the enrichment of noncoding DNMs that are likely functional in specific genes. Among the noncoding DNMs, we further classified 5,343 mutations in proband trios and 3,424 in unaffected sibling trios as likely functional, based on a CADD score ≥15, which account for the top ∼1.8% of variants. The distribution of noncoding DNMs is shown in **Fig 2**. The average number of noncoding DNMs with a CADD ≥ 15 is 1.52 per proband and 1.56 per unaffected sibling. We further replicated our pipeline in SSC, identifying 209,396 noncoding DNMs in 2274 affected offspring trios and 171,258 in 1835 unaffected sibling trios. The average of noncoding DNMs is 92.1 per proband and 93.3 per unaffected sibling. After filtering CADD score ≥ 15, we identified an average of 1.60 noncoding DNMs per ASD case and 1.59 per unaffected siblings in the final call set. The distribution of noncoding DNMs for SSC shown in **Fig S2**.

Case-control test was used to evaluate the burden of genic noncoding DNMs between ASD cases and unaffected sibling controls. We identified three genes with p <0.05 (DNM count ≥ 3, fold change>1, CADD score ≥15), including *TSHZ2*, *CTNNA2*, and *BCAS3*. *SCN2A* with 5 noncoding DNMs in ASD cases and none in unaffected siblings, with a p-value of 0.089. Additionally, we applied FATHMM-XF to annotate each SNV DNMs and identified four variants in *SCN2A* with FATHMM-XF scores greater than 0.5 in ASD cases, while none were observed in controls (**Table S4**). Among these, three had CADD scores ≥15, and one had a CADD score of 14.04 (**Table S5**). In SSC, *only SOBP* with genic noncoding DNMs reached suggested significance threshold (p <0.05, fold change>1, and DNM count ≥ 3).

We further evaluated the genes significantly enriched for noncoding DNMs in genic regions by comparing the observed gene-wise noncoding DNM count to that of expected (case-only test), and p value is calculated by a one tailed Poisson test. We identified 21 genes with suggestive evidence (p<0.05 by case-only test, CADD score ≥15, DNM count ≥3) by point-based test in genic regions. For further replication, we examined the genes in the SSC cohort, and found that 12 genes were enriched for noncoding DNMs (p<0.05 by case-only test, CADD score ≥15, DNM count ≥3). However, we did not identify overlapping genes that were enriched for noncoding DNMs in both cohorts with suggestive significance (p<0.05).

Finally, by combining evidence from case-only tests for both coding and genic noncoding DNMs using point-based test in genic regions, we identified 89 genes enriched for DNMs in SPARK (p<0.05 by case-only test). Notably, *SCN2A*, with 11 coding DNMs and 5 noncoding DNMs in probands and none in unaffected siblings (**Table 3**), shows the most significant DNM burden (p =4.15×10^−13^ by case-only test). Additionally, we estimated an odds ratio of 14.6 (p = 0.0045) for *SCN2A* coding DNMs and 6.97 (p = 0.0862) for noncoding DNMs.

**Table 3.**
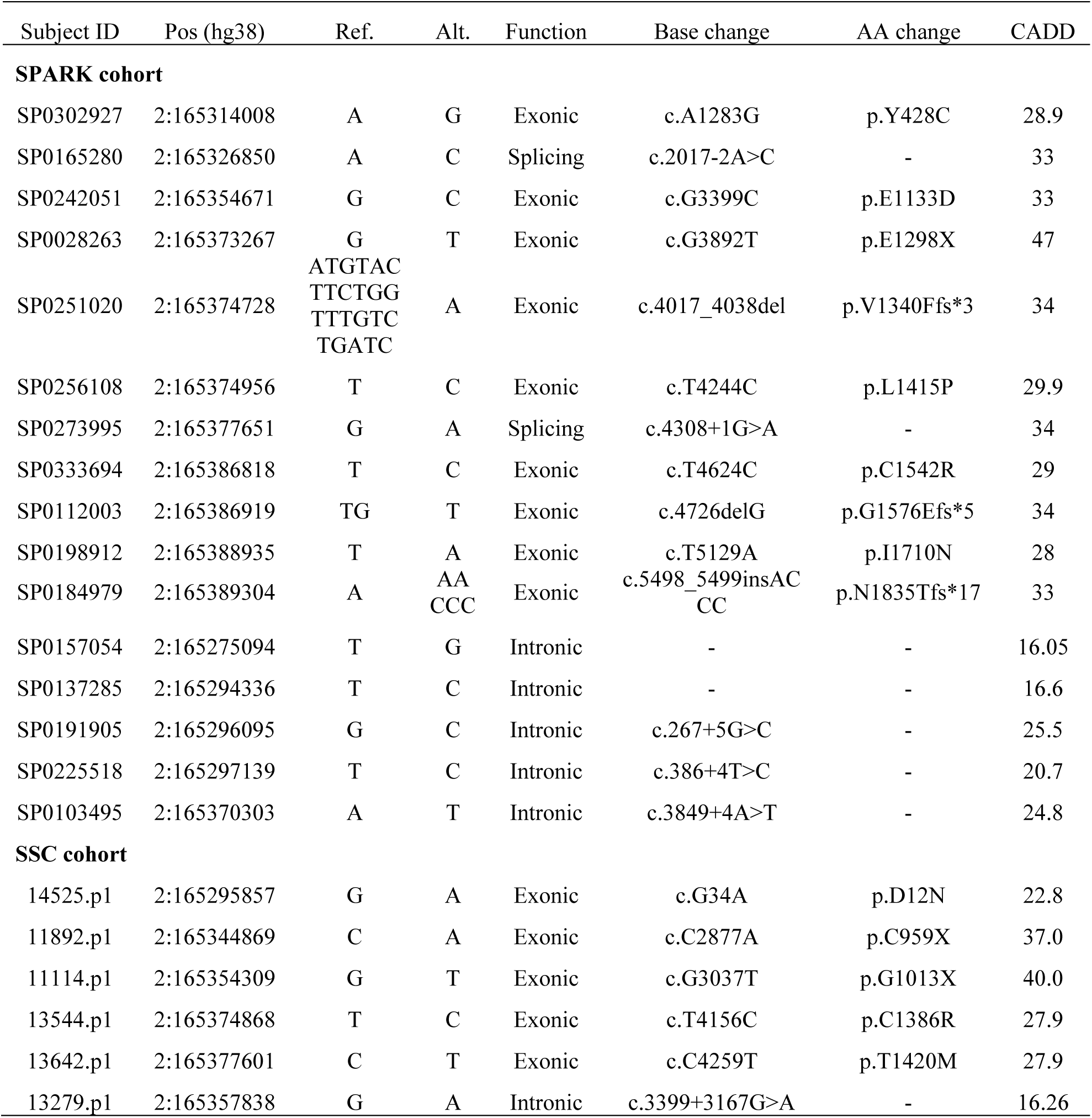
A list of DNMs in *SCN2A* from whole-genome sequencing of the SPARK cohort.

In the SSC cohort, 65 genes had an excess of DNMs (p<0.05 by case-only test). Among them, *SCN2A* had one noncoding DNM in probands and none in unaffected sibling control in SSC. Furthermore, the overlapping genes with suggestive significance (p<0.05) between SPARK and SSC cohorts include *SCN2A*, *DNMT3A*, *ARID1B*, *GRIN2B*, and *KDM6B.* The top genes in SPARK and SSC cohorts were showed in **Table 4**.

**Table 4.** Top genes with DNMs in the SPARK and SSC cohorts.

| Genes | Count of coding DNMs in cases | Count of coding DNMs in controls | Expected coding DNMs in cases | Count of noncoding DNMs with CADD $\geq 15$ in cases | Count of noncoding DNMs with CADD $\geq 15$ in controls | Expected noncoding DNMs with CADD $\geq 15$ in case | Case-only p value for coding DNMs | Case-only p value for noncoding DNMs | Combined p values for both coding and noncoding DNMs | SFARI gene score |
| --- | --- | --- | --- | --- | --- | --- | --- | --- | --- | --- |
| **Top 10 genes in SPARK cohort** |  |  |  |  |  |  |  |  |  |  |
| *SCN2A* | 11 | 0 | 0.55 | 5 | 0 | 0.66 | $2.06 \times 10^{-11}$ | $6.12 \times 10^{-4}$ | $4.15 \times 10^{-13}$ | 1 |
| *CCDC168* | 4 | 0 | 0.05 | 0 | 0 | 0.001 | $3.49 \times 10^{-7}$ | 1 | $5.53 \times 10^{-6}$ | - |
| *PIEZO1* | 3 | 0 | 0.11 | 1 | 0 | 0.02 | $2.13 \times 10^{-4}$ | 0.02 | $4.56 \times 10^{-5}$ | - |
| *PTEN* | 4 | 0 | 0.10 | 0 | 0 | 0.46 | $4.34 \times 10^{-6}$ | 1 | $5.79 \times 10^{-5}$ | 1 |
| *C16orf96* | 3 | 0 | 0.04 | 0 | 0 | 0.003 | $7.30 \times 10^{-6}$ | 1 | $9.37 \times 10^{-5}$ | - |
| *BRD4* | 6 | 0 | 0.47 | 0 | 0 | 0.40 | $9.95 \times 10^{-6}$ | 1 | $1.25 \times 10^{-4}$ | 2 |
| *ADNP* | 5 | 0 | 0.29 | 0 | 0 | 0.25 | $1.42 \times 10^{-5}$ | 1 | $1.72 \times 10^{-4}$ | 1 |
| *DDX3X* | 3 | 0 | 0.21 | 2 | 0 | 0.27 | $1.39 \times 10^{-3}$ | 0.03 | $4.70 \times 10^{-4}$ | 1 |
| *KDM6B* | 5 | 0 | 0.63 | 2 | 0 | 0.55 | $4.91 \times 10^{-4}$ | 0.11 | $5.60 \times 10^{-4}$ | 1 |
| *SOX1* | 4 | 1 | 0.21 | 0 | 0 | 0.05 | $7.46 \times 10^{-5}$ | 1 | $7.84 \times 10^{-4}$ | - |
| **Top 10 genes in SSC cohort** |  |  |  |  |  |  |  |  |  |  |
| *FLG2* | 8 | 5 | 0.58 | 0 | 0 | 0.01 | $7.05 \times 10^{-9}$ | 1 | $1.39 \times 10^{-7}$ | - |
| *CHD8* | 7 | 0 | 0.67 | 0 | 0 | 0.39 | $4.02 \times 10^{-7}$ | 1 | $6.32 \times 10^{-6}$ | 1 |
| *AHNAK2* | 10 | 5 | 1.99 | 0 | 0 | 0.05 | $1.11 \times 10^{-6}$ | 1 | $1.63 \times 10^{-5}$ | - |
| *SYNGAP1* | 5 | 1 | 0.48 | 1 | 2 | 0.64 | $1.94 \times 10^{-5}$ | 0.34 | $8.48 \times 10^{-5}$ | 1 |
| *AMZ1* | 4 | 1 | 0.19 | 0 | 0 | 0.07 | $9.49 \times 10^{-6}$ | 1 | $1.19 \times 10^{-4}$ | - |
| *SCN2A* | 5 | 0 | 0.55 | 1 | 0 | 0.66 | $3.53 \times 10^{-5}$ | 0.35 | $1.52 \times 10^{-4}$ | 1 |
| *PLIN4* | 5 | 4 | 0.51 | 0 | 0 | 0.02 | $2.45 \times 10^{-5}$ | 1 | $2.85 \times 10^{-4}$ | - |
| *WDFY4* | 3 | 0 | 0.09 | 0 | 0 | 0.06 | $3.15 \times 10^{-5}$ | 1 | $3.58 \times 10^{-4}$ | 2 |
| *CDC42BPB* | 3 | 1 | 0.62 | 2 | 0 | 0.18 | $8.06 \times 10^{-3}$ | 0.01 | $5.49 \times 10^{-4}$ | 2 |
| *ALMS1* | 5 | 1 | 1.03 | 1 | 1 | 0.19 | $6.38 \times 10^{-4}$ | 0.11 | $7.65 \times 10^{-4}$ | - |

| **Suggestive significant (p<0.05) in both cohorts (SPARK/SSC)** |  |  |  |  |  |  |  |  |  |  |
| --- | --- | --- | --- | --- | --- | --- | --- | --- | --- | --- |
| *SCN2A* | 11/5 | 0/0 | 0.55/0.36 | 5/1 | 0/0 | 0.66/0.43 | $2.60 \times 10^{-14}$ | $2.02 \times 10^{-3}$ | $2.41 \times 10^{-15}$ | 1 |
| *KDM6B* | 5/3 | 0/0 | 0.63/0.41 | 2/1 | 0/1 | 0.55/0.35 | $5.52 \times 10^{-5}$ | 0.14 | $1.23 \times 10^{-4}$ | 1 |
| *GRIN2B* | 5/3 | 0/0 | 0.54/0.35 | 2/0 | 0/1 | 2.81/1.82 | $1.91 \times 10^{-5}$ | 0.97 | $6.47 \times 10^{-4}$ | 1 |
| *DNMT3A* | 3/3 | 0/0 | 0.35/0.23 | 0/2 | 1/0 | 1.35/0.87 | $1.19 \times 10^{-4}$ | 0.55 | $1.14 \times 10^{-3}$ | 1 |
| *ARID1B* | 4/4 | 0/0 | 0.73/0.47 | 1/1 | 1/2 | 2.11/1.37 | $1.16 \times 10^{-4}$ | 0.93 | $2.66 \times 10^{-3}$ | 1 |

- 2 The results ranked by case-only p value for coding DNMs. Case-only p value is calculated as Poisson distribution using counts of - 3 observed/expected DNMs for each gene (case-only test). The combined p value for both coding and noncoding DNMs is calculated by Fisher - 4 combined test.

In addition to point-based test, we also used a segment-based test to evaluate the observed number of noncoding DNMs in genic region over expectation (case-only test). We identified 627 genes exhibiting excesses of noncoding DNMs in genic regions (p <0.05, DNM count ≥3; **Table S6**). Within this group, four genes (*CSMD1*, *WWOX*, *RBFOX1*, and *CDH13*) achieved Bonferroni corrected significance (p < 2.5×10^−6^ by case-only test). Moreover, our results show that the enrichment of noncoding DNMs in *CSMD1, WWOX, RBFOX1, and CDH13* remain statistically significant under FDR correction at a 5% threshold. We examined SSC for further replication and found that 535 genes are enriched for noncoding DNMs (*p* <0.05, DNM count ≥3) and four of these genes (*CSMD1*, *CDH13*, *RBFOX1*, *SGCZ*) met Bonferroni corrected significance (Bonferroni p <2.5×10^-6^). Therefore, *CDH13*, *CSMD1*, and *RBFOX1* reached exome-wide statistical significance across both cohorts (**Table 5**). However, in point-based test of noncoding DNMs with CADD>15, *CDH13*, *CSMD1*, and *RBFOX1* had 2, 3, and 19 observed noncoding DNMs in ASD cases, respectively. Therefore, while these genes tend to have more DNMs over expectation, likely due to being located at fragile genomic region, these DNMs are not predicted to be functionally important by CADD scores.

**Table 5.** Top genes with noncoding DNMs in the SPARK cohort, with validation in the SSC cohort, using point-based and segment-based statistical tests.

| Genes | SPARK cohort |  |  | SSC cohort |  |  | SFARI gene score |
| --- | --- | --- | --- | --- | --- | --- | --- |
|  | Count of noncoding DNMs in cases | Expected noncoding DNMs in cases | Case-only p value for noncoding DNMs | Count of noncoding DNMs in cases | Expected noncoding DNMs in cases | Case-only p value for noncoding DNMs |  |
| Point-based test: noncoding mutations with a CADD score ≥15 |  |  |  |  |  |  |  |
| Known ASD risk genes (*Zhou et al.*) |  |  |  |  |  |  |  |
| *SCN2A* | 5 | 0.66 | 6.12×10<sup>-4</sup> | 1 | 0.43 | 0.35 | 1 |
| *ZEB2* | 10 | 4.89 | 2.81×10<sup>-2</sup> | 4 | 3.17 | 0.39 | - |
| *AGMO* | 4 | 1.17 | 3.09×10<sup>-2</sup> | 1 | 0.76 | 0.53 | 2 |
| Newly found candidate ASD risk genes |  |  |  |  |  |  |  |
| *TSHZ2* | 9 | 2.96 | 3.51×10<sup>-3</sup> | 4 | 1.92 | 0.13 | - |
| *NELL1* | 7 | 2.10 | 5.88×10<sup>-3</sup> | 1 | 1.36 | 0.74 | - |
| Segment-based test: Gnocchi genome constraint in 1kb genomic segments |  |  |  |  |  |  |  |
| Known ASD risk genes (*Zhou et al.*) |  |  |  |  |  |  |  |
| *MAGI2* | 126 | 100.86 | 0.0087 | 86 | 64.41 | 0.0059 | - |
| *GRIN2A* | 48 | 33.50 | 0.0106 | 30 | 21.39 | 0.045 | 1 |
| Newly found candidate ASD risk genes (p<2.5×10<sup>-6</sup>) |  |  |  |  |  |  |  |
| *CSMD1* | 142 | 69.10 | 1.12×10<sup>-14</sup> | 104 | 44.13 | 2.98×10<sup>-19</sup> | 2 |
| *RBFOX1* | 208 | 120.15 | 2.44×10<sup>-13</sup> | 124 | 76.74 | 4.38×10<sup>-7</sup> | 2 |
| *CDH13* | 146 | 94.35 | 5.07×10<sup>-7</sup> | 110 | 60.26 | 5.86×10<sup>-9</sup> | 2 |

Case-only p value is calculated as Poisson distribution using counts of observed/expected DNMs for each gene (case-only test). SFARI gene score takes into account all available evidence supporting a gene's relevance to ASD risk and places each gene into a category reflecting the overall strength of that evidence. SFARI gene score rank genes into four categories, including syndromic (mutations that are associated with a substantial degree of increased risk and consistently linked to additional characteristics not required for an ASD diagnosis), category 1 (each of these genes has been clearly implicated in ASD, typically by the presence of at least three *de novo* likely-gene-disrupting mutations being reported in the literature), 2 (genes with two reported *de novo* likely-gene-disrupting mutations.), and 3 (genes with a single reported *de novo* likely-gene-disrupting mutation)

Next, we examined noncoding DNMs with CADD ≥15 by point-based test in intergenic regions. For this analysis, we assigned DNMs to its closest gene. We identified 54 genes enriched for noncoding DNMs in SPARK and 31 genes enriched for noncoding DNMs in SSC (p<0.05 by case-only test, CADD score ≥15, DNM count ≥3). Of which, *PRKD1* and *CHD8* met statistical significance in both SPARK and SSC (p < 2.5×10^−6^; **Table S7**). We further estimate gene-based noncoding DNMs by segment-based test in intergenic region, and only *CSMD1* reached statistically significant (p=2.79×10^−7^) in SPARK and marginally significant (p=5.47×10^−4^) in SSC (**Table S8**).

### Examination of noncoding DNMs in known ASD risk genes

To evaluate the contribution of noncoding DNMs within established ASD risk genes, we examined 618 well-documented dominant ASD genes. Among these, 22 genes (e.g., *SCN2A*, *NRXN3*, and *MEIS2*) that harbored three or more noncoding DNMs had higher enrichment in probands compared to unaffected siblings (CADD score ≥15, DNM count ≥3; **Table S9**), including *SCN2A*, *NRXN1*, *BCL11A*, *CASZ1*, *MEIS2*, *PBX1*, *EBF3*, *MEF2C*, *FOXP2*, *HDAC4*, *CUX2*, *SOX5*, *SATB1*, *EPB41L1*, *ZEB2*, *GLI3*, *NRXN3*, *RARB*, *MAGI2*, *CAMTA1*, *MACF1*, and

*TRPM3*. Remarkably, three out of these 22 genes—*SCN2A*, *HDAC4*, and *ZEB2*—are enriched for noncoding DNMs (p <0.05 by case-only test). For further replication, we examined the SSC cohort. This analysis revealed that, among the three genes (*SCN2A*, *HDAC4*, and *ZEB2*) identified with a higher burden of noncoding DNMs in SPARK, two genes (*SCN2A* and *ZEB2)* showed a higher burden in probands compared to unaffected siblings in SSC, though not reaching statistical significance.

When we applied a segment-based test to evaluate the excesses of noncoding DNMs for 618 known ASD risk genes, 23 genes are enriched for noncoding DNMs in SPARK and 22 genes are enriched in SSC (p<0.05 by case-only test). Moreover, the overlapping genes that are enriched for noncoding DNMs between SPARK and SSC cohorts include *MAGI2* and *GRIN2A* (p<0.05 by case-only test, **Table 5; Table S10**).

### Gene set analysis by case vs sibling control comparisons

We next characterized the enrichment of three gene sets with DNMs contributing to ASD risk in probands compared to unaffected trios. These include 3054 constrained genes (pLI>0.9), 1339 known NDD genes from Developmental Disorders Genotype-to-Phenotype database (DDG2P)^36^, and 618 known dominant ASD genes in Zhou et al^22^). Constrained genes (pLI>0.9) as a group are enriched for coding DNMs in cases than sibling controls (fold change=1.14, p =0.018 for SPARK, fold change=1.44, p =1.21×10^−7^ for SSC; **Table 6**). Moreover, the set of 618 known ASD genes had higher enrichment in both cohorts (fold change=1.51, p =1.13×10^−5^ for SPARK, fold change=1.86, p =2.06×10^−9^ for SSC). However, these three gene sets (LoF constrained, NDD, and known ASD genes) do not showed higher burden of noncoding DNMs in cases versus sibling controls (CADD≥15). Additionally, CWAS tests showed that noncoding variants are identified as enriched with regulatory elements, including enhancer and TFBS (case burden relative risk >1, p<0.05; **Table S11 and Fig S3**).

**Table 6.** Enrichment of DNMs in ASD cases across gene sets by case/control comparisons.

| Gene sets | SPARK cohort |  |  | SSC cohort |  |  |
| --- | --- | --- | --- | --- | --- | --- |
|  | Case/control<br>(# subjects) | Fold change | P value | Case/control<br>(# subjects) | Fold<br>change | P value |
| **Genes with coding DNMs** |  |  |  |  |  |  |
| All genes | 2484/1528 | 1.05 | 0.225 | 1717/1330 | 1.17 | 0.015 |
| LoF Constrained genes | 1000/567 | 1.14 | 0.018 | 726/451 | 1.44 | $1.21 \times 10^{-7}$ |
| NDD genes | 420/251 | 1.05 | 0.299 | 336/211 | 1.33 | 0.001 |
| Known ASD genes | 378/162 | 1.51 | $1.13 \times 10^{-5}$ | 303/140 | 1.86 | $2.06 \times 10^{-9}$ |
| **Genes with noncoding DNMs (point-based test with CADD>15)** |  |  |  |  |  |  |
| All genes | 2014/1297 | 0.93 | 0.922 | 1358/1093 | 1.01 | 0.472 |
| Constrained genes | 969/623 | 0.96 | 0.766 | 633/546 | 0.91 | 0.917 |
| NDD genes | 280/179 | 0.97 | 0.626 | 173/182 | 0.75 | 0.996 |
| Known ASD genes | 293/166 | 1.11 | 0.163 | 177/164 | 0.86 | 0.917 |
| **Genes with noncoding DNMs (segment-based test)** |  |  |  |  |  |  |
| All genes | 3508/2188 | 1 | 1 | 2274/1835 | 1 | 1 |
| Constrained genes<br>(231,967 1kb windows) | 3501/2183 | 1.15 | 0.516 | 2271/1828 | 2.90 | 0.098 |
| NDD genes<br>(64,749 1kb windows) | 2955/1872 | 0.90 | 0.918 | 1885/1554 | 0.88 | 0.944 |
| ASD known risk genes<br>(47,389 1kb windows) | 2662/1663 | 0.99 | 0.553 | 1738/1391 | 1.04 | 0.333 |
| **Genes with noncoding DNMs (segment-based test with Gnocchi score $\geq 4$)** |  |  |  |  |  |  |
| All genes<br>(12,792 1kb windows) | 1195/747 | 1.00 | 0.535 | 762/612 | 1.01 | 0.470 |
| Constrained genes<br>(2,993 1kb windows) | 351/231 | 0.94 | 0.763 | 233/180 | 1.05 | 0.341 |
| NDD genes<br>(1,198 1kb windows) | 141/84 | 1.05 | 0.395 | 90/85 | 0.85 | 0.873 |
| ASD known risk genes<br>(767 1kb windows) | 92/54 | 1.06 | 0.394 | 65/54 | 0.97 | 0.601 |

We defined 3 gene sets, including 3054 LoF constrained genes ( $pLI > 0.9$ ), 1339 known NDD genes from Developmental Disorders Genotype-to-Phenotype database (DDG2P), and 618 known dominant ASD genes. Chen et al.<sup>33</sup> built a genome-wide genomic constraint map (Gnocchi score) per 1-kb genomic windows by utilizing the Genome Aggregation Database (gnomAD V3.1.2), and highly constrained noncoding 1kb regions based on Gnocchi $> 4$ . Fold change calculated as the fraction of ASD cases with noncoding DNM compared to unaffected siblings (case-control test), and the p value is calculated by Fisher exact test.

We further evaluate the enrichment of noncoding DNMs identified by segment-based test. We failed to find any significant enrichment of noncoding DNMs in cases over sibling controls in any of the gene sets. Additionally, when we focus on 1kb segments with Gnocchi score>4, we still do not see increased burden of noncoding DNMs in cases over controls (**Table 6**).

### Function prediction of noncoding DNMs in SCN2A, ZEB2, and AGMO

Three noncoding DNMs located in *SCN2A* (chr2:165296095, G>C; chr2:165297139, T>C; and chr2:165370303, A>T) are situated less than 20 base pairs (bp) away from an exon and are highly conserved (**Fig 4**). The probability that the position 2:165296095 (5 bp away from an exon) being splice donor loss is 0.98 (delta score ≥ 0.8 is high precision). Likewise, the probability that the position 2:165370303 (4 bp away from exon) being splice donor loss is 0.81. In addition, the noncoding DNM found at position chr2:165294336 (T>C) of *SCN2A* is located within a candidate cis-Regulatory Element (cCRE) with a proximal enhancer like signature.

**Fig 4.**
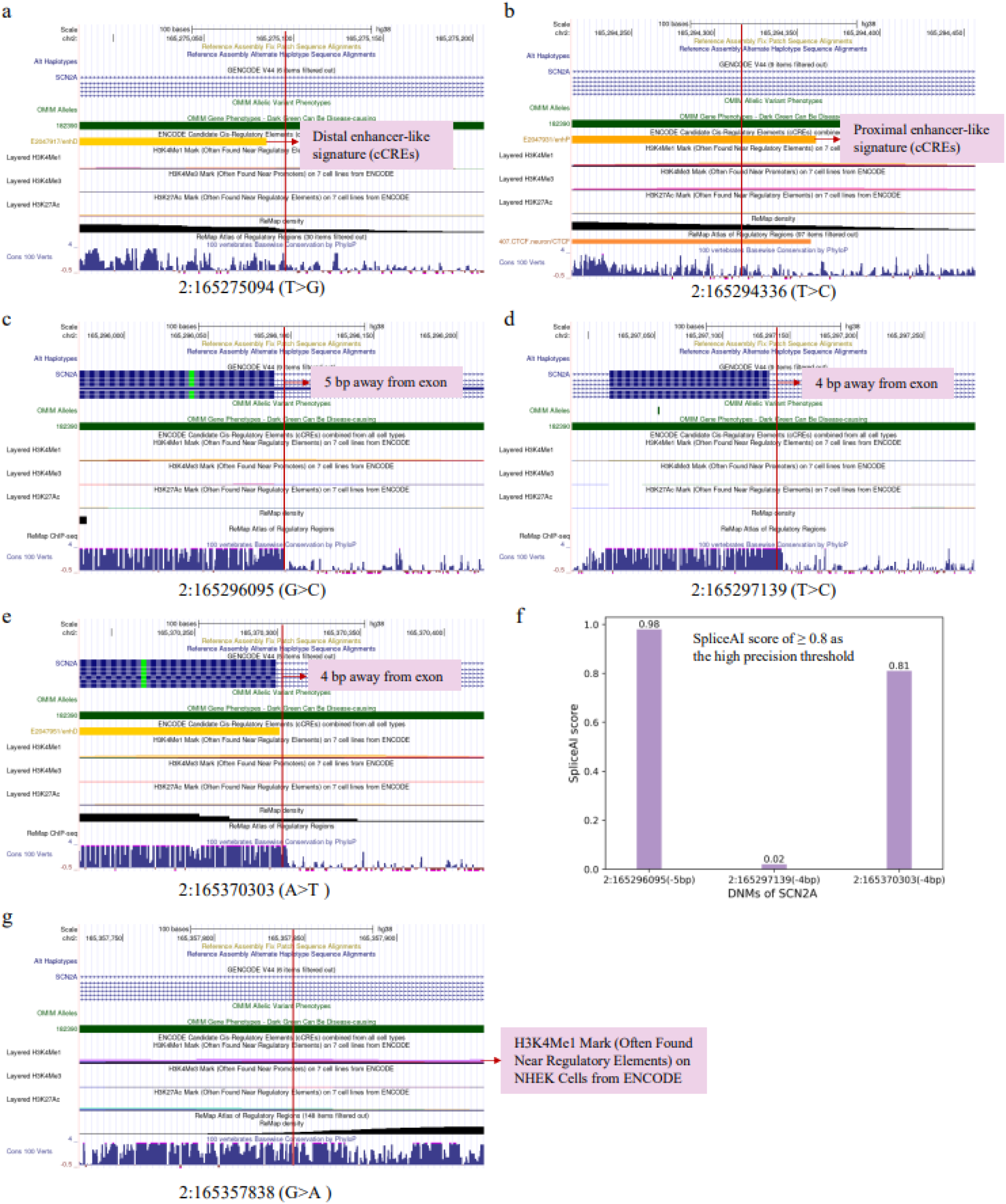
Mutation of the noncoding DNMs for the *SCN2A* in SPARK and SSC. **(a)** mutation of chr2:165275094 (T>G) in SPARK. **(b)** mutation of chr2:165294336 (T>C) in SPARK. **(c)** mutation of chr2:165296095 (G>C) in SPARK. **(d)** mutation of chr2:165297139 (T>C) in SPARK. **(e)** mutation of chr2:165370303 (A>T) in SPARK. **(f)** probability of the mutations situated less than 20 base pairs (bp) distant from the exon being splice-altering in SPARK. (**g**) mutation of 2:165357838 (G>A) in SSC. In SAPRK, mutation of chr2:165294336 (T>C) likely a candidate cis-Regulatory Element (cCRE) with a proximal enhancer like signature. Three noncoding DNMs (2:165296095, 2:165297139, and 2:165370303) located in *SCN2A* are situated less than 20 bp away from an exon, and two noncoding DNMs (2:165296095 and 2:165370303) likely being splice-altering (SpliceAI score >0.8).

In the *ZEB2* gene, 10 noncoding DNMs were identified in probands and 3 in unaffected sibling controls (2.07-fold). Two of the 10 noncoding DNMs in probands were identified as cCREs with a distal enhancer-like signature. One noncoding DNM, located at position chr2:144404388 (T>C), showed higher enrichment of the H3K27Ac histone mark on HSMM Cells, as determined by a ChIP-seq assay (**Fig S4**). Another one, located at position chr2:144504145 (G>A), exhibited higher enrichment of the H3K4Me1 histone mark on K562 Cells. Additionally, one noncoding DNM (chr2: 144396373, G>C) is situated 39 bp away from the exon, while this mutation less likely being a splice site (delta score=0).

The *AGMO* gene harbored 4 noncoding DNMs in ASD cases. One noncoding DNM, located at position chr7:15556988 (T>C), shows characteristic of a distal enhancer-like signature and higher enrichment of the H3K4Me1 histone mark on HUVEC cells (**Fig S5**). Additionally, mutation in chr7:15502483 (G>A) shows slightly higher enrichment of the H3K4Me1 histone mark on HUVEC cells.

## Discussion

In the current study, we developed a custom pipeline to identify both coding and noncoding DNMs across 3357 families (12,411 individuals) with whole genome sequencing data from the SPARK cohort, while examining SSC with 6383 individuals from 2274 families to replicate the results. For coding DNMs, *SCN2A* reached exome-wide significance in SPARK. The 618 known dominant ASD genes as a group are strongly enriched for coding DNMs in cases than sibling controls. For noncoding DNMs, the point-based test identified *SCN2A* as marginally significant in SPARK, yet segment-based test identified *CSMD1*, *RBFOX1* and *CDH13* as exome-wide significant. We did not identify significant enrichment of noncoding DNMs (in all 1kb segments or those with Gnocchi>4) in the 618 known ASD genes as a group in cases than sibling controls. When combining evidence from both coding and noncoding DNMs, we found that *SCN2A* with 11 coding and 5 noncoding DNMs exhibited the strongest significance.

Previous Studies have reported an average of 60-100 DNMs per genome per generation^37–39^. Our study identified an average of ∼90 DNMs per child in the final call set, which is consistent with previous studies. Furthermore, our analysis identified an average of 1.3 coding DNMs per child in SSC, which closely aligns with previous findings (e.g., Werling et al.^29^ reported approximately 1.2 coding DNMs per child). An et al. reported a total of 255,106 DNMs across 1,902 families, with 61.5 de novo single nucleotide variants (SNVs) and 5.6 de novo indels per child^40^. These numbers are lower than our current study’s estimate, which may reflect differences in variant filtering thresholds, genomic regions considered, or updates in gene models and annotations since the original publications.

ASD is characterized by its clinical and etiological heterogeneity, which makes it difficult to elucidate the neurobiological mechanisms underlying its pathogenesis. Recently, DNMs have been recognized as strong source of genetic causality, and the characterization of DNMs allows additional ASD risk genes to be identified. DNMs in non-coding regions have become of interest in recent years. Previous whole exome sequencing studies were unable to detect these variants due to the lack of coverage and sequencing depth across non-coding regions. However, there is evidence that ASD genes harbor hotspots of hypermutability in non-coding regions and that deleterious mutations across them are subjected to strong negative selection just like the loss of function mutations located in the coding region^13^. Werling et al. present an analytical framework to evaluate rare and *de novo* noncoding mutations from whole genome sequencing of 519 ASD families unable to demonstrate a rare noncoding variant contribution to ASD risk, but found that noncoding *de novo* indels category showed a greater number of nominally significant results than expected^29^. The authors^29^ demonstrated that the contribution of *de novo* noncoding variation is probably modest compared to *de novo* coding variants. Although whole genome sequencing now allows the identification of noncoding DNMs in affected probands, their contribution to risk remains relatively unexplored and demands further investigation. Our study suggested that noncoding DNMs of *SCN2A* were associated with ASD risk.

The *SCN2A* gene encodes the voltage-gated NA^+^ channel Nav1.2, one of the major neuronal sodium channels that play a role in the initiation and conduction of action potentials^41^.

Pathogenic mutations in *SCN2A*, have been associated with a spectrum of epilepsies and neurodevelopmental disorders^42–46^. Moreover, multiple studies have confirmed the contribution of *de novo SCN2A* mutations to the risk for ASD. Stephan et al. using whole-exome sequencing of 928 individuals, including 200 phenotypically discordant sibling pairs confirmed that *de novo* coding mutations across *SCN2A* were associated with ASD risk^46^ . Our discovery of coding DNMs events in *SCN2A* related to the risk of ASD are supporting previous studies. Nevertheless, the estimated odds ratio for noncoding DNMs in *SCN2A* is approximately half that of its coding DNMs in our cohort, consistent with previous findings that de novo protein-truncating variants (PTVs) and missense mutations carry stronger ASD risk than noncoding mutations^40^.

Intriguingly, three noncoding DNMs events observed in *SCN2A* are located within 20 base pairs of exons, further implicating them in ASD risk. Notably, two of these noncoding DNMs demonstrated a high likelihood of affecting splice donor/loss sites, potentially leading to significant alterations in gene expression. Also, one noncoding DNM in *SCN2A* displays characteristic of a proximal enhancer like signature. This suggests a critical role for both coding and proximal noncoding regions of *SCN2A* in ASD pathogenesis, highlighting the intricate interplay between genetic variants and their potential impact on splicing and gene function.

Nevertheless, none of the five probands carrying the noncoding DNMs in *SCN2A* were also found to harbor a coding DNM in the same gene. Therefore, based on current annotations and predicted regulatory impact (CADD≥15, disrupt splice sites, cCRE overlap), we consider the *SCN2A* noncoding DNMs in these individuals to be the plausible candidate causative variants. Further experimental studies are still needed to validate the contribution of these noncoding variants to ASD risk.

*CHD8* encodes chromodomain helicase DNA-binding protein 8 and its mutation is a highly penetrant risk factor for ASD^21,47^. While DNMs of *CHD8* are associated with ASD risk in the SSC cohort, but not in the SPARK cohort. One possible explanation may be the different sample characteristics and family fractures. In SSC cohort, over 80% family types are quartet families and around 20% are trios. While in SPARK cohort, 65% family types are quartet families and 33% are trios. Additionally, the stochastic nature of rare mutations may also explain the general lack of DNMs in *CHD8* in the SPARK cohort.

In comparison with previous large-scale studies such as Satterstrom et al.^21^ and Fu et al.^48^, which identified 102 and 373 ASD risk genes respectively using WES and WGS data focused on coding variation, our study highlights both overlapping and distinct gene sets. While we replicate several well-established coding ASD genes (e.g., *SCN2A*, *ADNP*, *PTEN*), we also identify novel candidates (*CSMD1*, *RBFOX1*, *CDH13*) driven by enrichment of noncoding DNMs. These differences can be attributed to variations in methodology, such as our use of a statistical framework described and validated by Samocha et al.^35^ versus the TADA framework^21,48^, as well as differences in sample composition and statistical thresholds. This underscores the importance of integrating findings from diverse analytic strategies to fully capture the genetic architecture of ASD.

Both our method and CWAS found that noncoding DNMs enriched in regulatory elements. These overlapping results support the robustness of the underlying biological signals and highlight the complementary nature of the two approaches. Besides, our method differs from CWAS in several key aspects. CWAS test for enrichment of DNMs across predefined functional categories (e.g., enhancers, TFBS), while our method assigns noncoding DNMs with predicted impact (e.g., CADD ≥15) to genes and tests for gene-level enrichment. Moreover, CWAS incorporates diverse annotations such as chromatin accessibility, conservation, and enhancer activity, aiming to capture the aggregate burden of mutations in biologically informed regions. Our method utilized functional prediction scores like CADD to prioritize likely deleterious noncoding DNMs, and further examined enrichment within LoF constrained, NDD, and known ASD genes. CWAS operates at the category level, providing insight into mutation enrichment in functional genomic elements. Our approach operates at the gene level, enabling direct prioritization of ASD candidate genes (e.g., *SCN2A*, *ZEB2*) with supporting evidence from both coding and noncoding DNMs. The advantage of our approach is its ability to directly implicate genes with functional evidence from both coding and noncoding variants, which facilitates biological interpretation and downstream validation. However, this focus on gene assignment may miss distal regulatory elements that act over long genomic distances. CWAS excels at capturing global burden in functional annotations, but its results are more difficult to connect to specific genes without additional mapping steps. Additionally, CWAS depends on accurate annotation of functional categories and large sample size, while our method relies more heavily on prediction accuracy for functional scores. In future studies, a hybrid framework that combines CWAS’s category-level sensitivity with our gene-centric focus may offer a more comprehensive view of noncoding DNM contribution to ASD risk.

### Limitations of the study

To our best knowledge, this study was among the first to evaluate the significance of non-coding DNMs implicated in the risk of ASD from whole genome sequencing data across more than 5000 families. Our findings also suggested the contribution of noncoding DNMs in known ASD risk genes, especially *SCN2A*. This study also has several limitations. The sample size with whole-genome sequencing data from SPARK is relatively small, and the size of the ASD cases was around double that of the unaffected sibling controls, which may hinder the discovery of ASD risk genes. Moreover, ASD can be diagnosed at different ages, and it is possible that some of the younger unaffected siblings may be diagnosed later, which could affect our results estimate. Therefore, future follow-up studies would be valuable in assessing the potential for later diagnoses. Furthermore, the probability of DNMs per gene may differ between coding and noncoding regions. To compute the expected number of noncoding DNMs, we assume that the mutation rate of the noncoding DNMs occurring within the same gene is comparable to that of coding DNMs^49,50^. Rodriguez-Galindo et al. demonstrated that the rate of generation of new genetic variants, the mutation rate, does not significantly vary between exons and adjacent introns when accounting for sequence context^51^. However, Sankar et al. revealed mutation rates in exons are 30%–60% higher than in noncoding DNA due to the relative overabundance of synonymous sites involved in CpG dinucleotides^52^. Studies also reported that mutation rate in non-coding regions is highly heterogeneous and can be affected by local sequence context as commonly modelled in gene constraint metrics as well as by a variety of genomic features at larger scales^53,54^. Therefore, we used two different methods (point-based and segment-based) to model the statistical significance of noncoding DNMs. In the near future, repeating these analyses with large-scale whole genome sequencing data, through combined analysis of coding and noncoding DNMs and structural variants, and incorporating inherited variants will aid in identifying additional ASD risk genes. Recent large-scale WGS studies of East Asian ASD families provide valuable opportunities for cross-ancestry comparison of coding and noncoding de novo variants^55^. While our study focuses on predominantly European ancestry populations from the SPARK and SSC cohorts, which limits the generalizability of our findings across global populations, integrating WGS datasets from diverse populations through meta-analysis may improve detection power and reveal ancestry-specific noncoding risk loci. Our pipeline is adaptable to different cohorts, it may require adjustments, such as background mutation rate and annotation frameworks, to identify population-specific regulatory elements. A broader population-based approach will be crucial in future efforts to fully elucidate the genetic architecture of ASD across different ancestries.

## Data Availability

All data produced are available online at https://www.sfari.org/resource/sfari-base.

## Acknowledgements

We are grateful to all of the families at the participating Simons Simplex Collection (SSC) sites, as well as the principal investigators (A. Beaudet, R. Bernier, J. Constantino, E. Cook, E. Fombonne, D. Geschwind, R. Goin-Kochel, E. Hanson, D. Grice, A. Klin, D. Ledbetter, C. Lord, C. Martin, D. Martin, R. Maxim, J. Miles, O. Ousley, K. Pelphrey, B. Peterson, J. Piggot, C. Saulnier, M. State, W. Stone, J. Sutcliffe, C. Walsh, Z. Warren, E. Wijsman). We are grateful to all of the families in SPARK, the SPARK clinical sites and SPARK staff. We appreciate obtaining access to genetic data on SFARI Base. Approved researchers can obtain the SSC population dataset described in this study by applying at https://base.sfari.org. Approved researchers can obtain the SPARK population dataset described in this study by applying at https://base.sfari.org. We appreciate obtaining access to recruit participants through SPARK research match on SFARI Base. We thank Dr. Yufeng Shen (Columbia University) for his insightful comments on the analysis strategies. The study is supported in part by a Foerderer grant, NIH grant HG013031 and the CHOP Research Institute.

## Author contributions

Kai Wang conceptualized and designed the study. Yuan Zhang drafted the manuscript. Yuan Zhang and Mian Umair Ahsan contributed to analysis and interpretation of data. Kai Wang was involved in study supervision. All authors contributed to the intellectual content, critical revisions to the drafts of the paper and approved the final version.

## STAR Methods

Key resources table

| REAGENT or RESOURCE | SOURCE | IDENTIFIER |
| --- | --- | --- |
| Deposited data |  |  |
| Simons Foundation Powering Autism Research for Knowledge (SPARK) | SFARI Base | https://www.sfari.org/resource/sfari-base/ |
| Simons Simplex Collection (SSC) | SFARI Base | https://www.sfari.org/resource/sfari-base/ |
| Software and algorithms |  |  |
| Python 3.10.11 | Python Software Foundation | https://www.python.org/ |
| R 4.3.3 | R Foundation | https://www.r-project.org/ |
| ANNOVAR | Wang et al<sup>56</sup> | https://annovar.openbioinformatics.org/en/latest/ |
| Bcftools 1.17 | Petr et al<sup>57</sup> | http://github.com/samtools/bcftools |
| Bedtools v2.31.0 | Quinlan et al<sup>58</sup> | https://github.com/arq5x/bedtools2 |
| gnomAD version 3.1.2 | Chen et al<sup>33</sup> | https://gnomad.broadinstitute.org |
| The UCSC Genome Browser | Nassar et al<sup>59</sup> | https://genome.ucsc.edu |
| SpliceAI | de Sainte Agathe et al<sup>60</sup> | https://spliceailookup.broadinstitute.org |
| CADD score | Philipp et al<sup>32</sup> | https://cadd.gs.washington.edu/ |
| FATHMM-XF | Rogers et al<sup>61</sup> | https://fathmm.biocompute.org.uk/fathmm-xf |
| CWAS-plus | Kim et al<sup>62</sup> | https://github.com/joonan-lab/cwas |

### Resource availability

#### Lead contact

Further information and requests should be directed to the lead contact, Kai Wang.

#### Materials availability

All relevant data are within the manuscript and supporting information files.

#### Data and code availability

This paper shares the original code at https://github.com/WGLab/ SPARK_de_novo_analysis_pipeline. Whole genome sequencing data was made available via SFARI and can be requested through SFARI Base (https://www.sfari.org/resource/sfari-base/). Any additional information required to the data reported in this paper is available from the lead contact upon request.

#### Experimental model and study participant details

This study uses whole genome sequencing data of 12,411 individuals from 3,357 families collected in SPARK to detect DNMs associated with ASD, while SSC with 6383 individuals from 2274 families to replicate the results. The procedure including sample selection and quality control of data are described in the method details section. All participants were recruited to SPARK under a centralized institutional review board (IRB) protocol (Western IRB Protocol no. 20151664). All participants signed the written informed consent.

## Method details

### Sample selection

The whole genome sequencing data of SPARK (v1.1, released January 26, 2023) was made available via Simons Foundation Autism Research Initiative (SFARI) and can be requested through SFARI Base (https://www.sfari.org/resource/sfari-base/). All participants were recruited to SPARK under a centralized institutional review board (IRB) protocol (Western IRB Protocol no. 20151664). Written informed consent was obtained from all legal guardians or parents for all participants aged 18 and younger, as well as for those aged 18 and older who have a legal guardian. Assent was also obtained from dependent participants aged 10 and older. In total 3385 families were selected from the SPARK cohort. We excluded 28 families with missing data on paternal and/or maternal whole genome sequencing, leaving 3357 families (12,411 individuals) for analysis. Genetically inferred race/ethnicity by multidimensional scaling (MDS) analysis indicate that ∼ 75% of the population is White (**Fig S1**), and self-reported race/ethnicity is provided in **Table S1**. Our study was approved by the Institutional Review Board of the Children’s Hospital of Philadelphia.

The SSC (v2, released January 26, 2023) cohort also represents an important data resource from the SFARI, to identify *de novo* genetic risk variants that contribute to the ASD^34^. Up to date, more than 2000 families with whole genome sequencing and clinical data have been collected. Our study included 6383 individuals (including 2274 affected probands and 1835 unaffected siblings) with whole genome sequencing from 2274 families to replicate results from the SPARK cohort. Probands were excluded who were younger than 4 years of age or older than 18. Informed consent was obtained at each data collection site included in the SSC.

### Variant calling

Sequencing and genotyping of all samples for SPARK and SSC were performed at New York Genome Center (NYGC). Alignment of reads to the human reference genome version GRCh38, duplicate read marking, and Base Quality Score Recalibration (BQSR) were performed using the standard pipeline from the Centers for Common Disease Genomics (CCDG). Variants calls for all samples from the SPARK and SSC cohorts are provided from NYGC.

DeepVariant gVCFs are provided for all samples enrolled in SPARK. DeepVariant version 1.3.0 was used to call SNVs and INDELs to produce sample-level gVCFs using the default WGS configuration profile (“--model_type=WGS”). All samples were then jointly called using GLnexus version 1.4.1 and the default DeepVariant WGS configuration (“--config DeepVariantWGS”). Genomic “chunks” were processed in parallel for computational feasibility, then subsequent chunks were combined to form project VCFs (pVCFs) by chromosome. Post-calling BCFtools (version 1.17) norm was used to left-align and normalize indels.

For SSC cohort, the SNVs and indels in families were called using four different callers: GATK HaplotypeCaller v.3.5.0, FreeBayes v1.1.0, Platypus v0.8.1, and Strelka2 v2.9.2. In addition, multi-nucleotide variants were called using FreeBayes and Platypus. Post-calling BCFtools (version 1.3.1) norm was used to left-align and normalize indels. They partitioned the genome into the high-quality regions, consisting of unique space as well as ancient repeats, and the recent repeat regions, which consisted of repeats <10% diverged from the consensus in RepeatMasker. Variants were only assessed in high quality portions of the genome and those in recent repeat regions were removed from the study. Our study selected project VCFs (pVCFs) generated by GATK HaplotypeCaller to replicate *de novo* analysis.

### DNMs detection

We developed a custom pipeline for DNM analysis. Candidate DNMs, defined as variants present in the offspring and absent in both parents, were identified from per-family VCFs generated by DeepVariant. Bcftools and Bedtools (v2.31.0) were used to further filter the candidate DNMs. The filtering criteria for DNMs as follows: 1) Allelic depth (AD) ≥ 6 in the offspring, 2) Genotype quality (GQ) ≥ 25 in the offspring and GQ ≥ 20 in the parents, 3) Read depth (DP) > 8 in the parents, 4) Annotated the candidate DNMs using a custom pipeline based on ANNOVAR^56^ (human genome hg38), and filtering the calls with an allele frequency (genomAD) < 0.1%, 5) Removed DNMs located in low mappability regions, 6) Filtering the fraction of reads supporting the alternate allele (AB.ALT) at 0.25∼0.75 in the offspring, 7) removed variants located in regions known to be difficult for variant calling (e.g., HLA gene and MUC gene), and 8) when an individual carries multiple DNMs within 100 bp in the same gene, only one variant with the most severe effects was included in the analysis. CADD was used to score the deleteriousness of noncoding mutations. To identify potentially pathogenic variants, we extracted noncoding DNMs with CADD score thresholds of 15, which rank among the top ∼1.8% of variants with the most severe predicted effect. Meanwhile, FATHMM-XF^61^ are available for all SNVs except for indels and sex chromosome, and prediction scores above 0.5 are predicted to be pathogenic. We further using FATHMM-XF scores for comparison. Pipelines for analysis were blind with respect to affected probands and unaffected siblings.

### Enrichment of coding DNMs

To account for family structure of quartets, we compared the burden of DNMs between affected probands and their unaffected siblings (case-control comparison). Fold enrichment was calculated as the ratio of DNMs in affected versus unaffected siblings, and Fisher’s exact test was used to assess whether the difference was statistically significant.

We further determined whether any individual genes carry an excess of DNMs. To compute the expected number of coding DNMs (including exonic and canonical splicing regions) in the cohort, we used pre-computed tabulation of the probability of DNMs arising in each gene based on RefSeq transcript definitions^35^. The number of expected coding DNMs equals to a gene’s inferred DNMs mutation rate multiply by the number of trios and by 2 (for the number of chromosomes for autosomes). Given that the number of DNMs per trio follows a Poisson distribution^63^, we use the Poisson test to evaluate the excesses of *de novo* events:

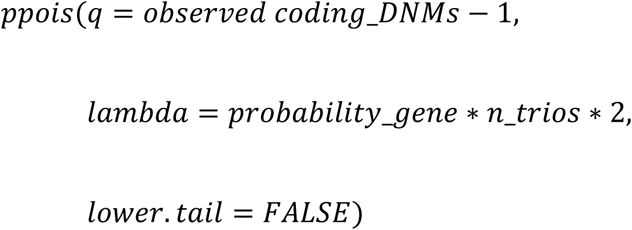

### Enrichment of noncoding DNMs

For noncoding DNMs, we used two methods to assess statistical significance: a point-based test that analyzes sites with a CADD score ≥15, and a segment-based test that uses genomic non-coding constraint of haploinsufficient variation (Gnocchi)^33^ constraint scores in 1kb genomic segments to infer the background mutation rates.

1. Point-based test: to compute the expected number of noncoding DNMs, we assumed that the probability of the noncoding mutations within a given gene aligns with that of coding DNMs. Rodriguez-Galindo et al. supported that the *de novo* mutation rate is similar in exons compared to introns in the germline, after accounting for trinucleotide sequence composition and an excess of nonsynonymous exonic variation arising from sampling bias^51^. Given that gene length is an obvious factor in a gene’s mutability (gene length showed high correlation (r = 0.880) with the probability of DNMs in each gene^35^), the expected number of noncoding DNMs is calculated as follows: multiply the gene’s DNM mutation rate by the number of trios, then by 2 (for autosomes), and finally by the ratio of the noncoding gene length to the coding gene length. Here, the noncoding variants are classified as those in genic (intronic, upstream, downstream, ncRNA exonic, ncRNA intronic, ncRNA splicing, UTR5, and UTR3) regions. We separately calculated statistical significance for intergenic regions by assigning noncoding variants to the nearest genes. CADD scores were available for all of the noncoding DNMs included in our analysis. Among the noncoding DNMs, we further classified mutations as severe if they had a CADD score greater than 15. Consequently, the noncoding gene length were calculated based on mutations with CADD score ≥15. Finally, we computed the p value for the observed number of noncoding DNMs compared to the expected number of noncoding DNMs

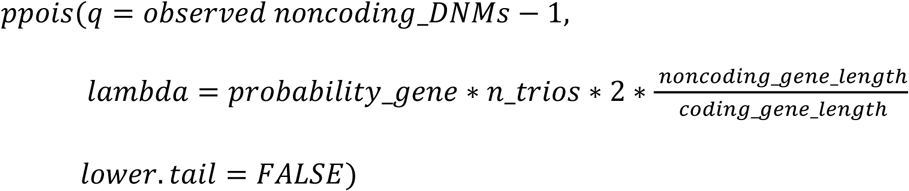
2. Segment-based test: Chen et al.^33^ built a genome-wide genomic constraint map (Gnocchi) per 1-kb genomic windows by utilizing the Genome Aggregation Database (gnomAD version 3.1.2). The authors derived Gnocchi score by comparing the observed mutation to an expectation from1,984,900 tiling 1kb genomic windows (passing all quality control checks) on autosomes. We annotated 1kb genomic windows and excluded variants located within coding and intergenic regions, leaving 862,475 1kb noncoding bins. We used Gnocchi genome constraint in 1-kb genomic segments to calculate the expected number of noncoding mutations in our cohorts as follows:

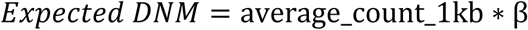

Where average count 1kb is the average number of DNM per 1kb, which is calculated as the average number of noncoding DNMs in a test dataset among 876,086 noncoding 1 kb genomic windows. β is inferred directly from Gnocchi calculation ^33^ which includes a score of the expected number of rare variants per 1kb genomic segments based on genomic contexts, and it is calculated as the ratio of the counts of the expected mutations in this 1kb bin divided by the average number of expected mutations in all 1kb bins in gnomAD.

We computed the total number of DNMs within genomic bins assigned to each gene, and calculates P value using a Poisson distribution based on expected and observed mutation counts per gene. To account for multiple comparisons, we applied both Bonferroni correction (p < 2.5 × 10⁻⁶ for ∼20,000 genes) and False Discovery Rate (FDR) correction using the Benjamini-Hochberg procedure. Genes passing FDR < 0.05 were considered statistically significant after controlling for the expected proportion of false discoveries.

To characterize the enrichment of groups of genes with DNMs contributing to ASD risk, we defined 3 gene sets, including 3054 LoF constrained genes (pLI>0.9), 1339 known NDD genes from Developmental Disorders Genotype-to-Phenotype database (DDG2P)^36^, and 618 known dominant ASD genes (**Table S2**). This set of 618 known ASD genes are compiled in Zhou et al^22^, and they encompass known NDD genes from DDG2P, high-confidence ASD genes collected by the SFARI, and dominant ASD genes included in the SPARK genes list. Additionally, for highly constrained noncoding 1kb regions based on Gnocchi threshold (for example, Gnocchi>4), we calculated the burden of genes with noncoding DNMs via case-sibling comparison with score over this threshold. Category-Wide Association Study (CWAS) is a data analytic tool to perform stringent association tests to find noncoding variants associated with autism disorder. Werling et al.^29^ first introduced CWAS analysis in *Nature Genetics* in 2018, and Kim et al.^64^ later enhanced the method to CWAS-plus in 2024, reducing computational time and memory usage. CWAS enhances noncoding variant analysis by integrating both WGS and user-provided functional data. In this study, we applied CWAS to perform a comparison with our method.

### Functional prediction of noncoding variants

Genome browser (https://genome.ucsc.edu) was used to visualize and browse noncoding DNMs with a CADD score ≥15, and candidate cis-regulatory elements (cCREs) derived from ENCODE^65^ integrated DNase-based sequencing (DNase-seq) and chromatin immunoprecipitation with sequencing (ChIP–seq) data. Genes with an pLI > 0.9 are defined as LoF constrained genes. We annotated potential splice site variants using SpliceAI^60^, which predicts splice site gain or loss events, and we set delta scores of ≥0.8 as the high precision threshold.

## Supporting information Document S1. Table S1–S11

**Table S1.** Self-reported race/ethnicity in SPARK

**Table S2:** Three gene sets (pLI >0.9, NDD, known ASD genes)

**Table S3.** Coding and noncoding DNMs (with CADD ≥15) of gene

**Table S4.** Noncoding single nucleotide variant (SNV) DNMs of genes based on CADD and FATHMM-XF scores

**Table S5.** A list of DNMs in SCN2A from whole-genome sequencing of the SPARK and SSC cohort, including CADD and FATHMM-XF scores

**Table S6.** Noncoding DNMs of genes based on segment-based test

**Table S7.** Noncoding DNMs (with CADD ≥15) of genes within intergenic regions

**Table S8.** Noncoding DNMs of known ASD risk genes based on segment-based test

**Table S9.** Coding and noncoding DNMs (with CADD ≥15) for 618known ASD risk genes

**Table S10.** Noncoding DNMs of genes for intergenic regions based on segment-based test

**Table S11.** Result of category-wide association tests from SPARK cohort

## Document S2. Figure S1–S5

**Fig S1.** Genetically inferred race/ethnicity by multidimensional scaling (MDS) analysis in SPARK

**Fig S2.** Distribution of DNMs in SSC

**Fig S3.** A category-wide association study (CWAS) analysis from SPARK cohort

**Fig S4.** Mutation of the noncoding DNMs for *ZEB2* in SPARK

**Fig S5.** Mutation of the noncoding DNMs for *AGMO* in SPARK

